# A low-cost molecular test for SARS-CoV-2 detection suitable for variant discrimination and community testing using saliva

**DOI:** 10.1101/2023.07.20.23292863

**Authors:** Sofia M. da Silva, Catarina Amaral, Cláudia Luís, Diana Grilo, Américo Duarte, Inês Morais, Gonçalo Afonso, Nuno Faria, Wilson Antunes, Inês Gomes, Raquel Sá-Leão, Maria Miragaia, Mónica Serrano, Catarina Pimentel

## Abstract

The gold standard for COVID-19 diagnostic testing relies on RNA extraction from naso/oropharyngeal swab followed by amplification through RT-PCR with fluorogenic probes. While the test is extremely sensitive and specific, its high cost and the potential discomfort associated with specimen collection make it suboptimal for public health screening purposes.

In this study, we developed an equally reliable, but cheaper and less invasive alternative test based on a one-step RT-PCR with the DNA-intercalating dye SYBR Green, which enables the detection of SARS-CoV-2 directly from saliva samples or RNA isolated from nasopharyngeal swabs. Importantly, we found that this type of testing can be fine-tuned to discriminate SARS-CoV-2 variants of concern.

The saliva RT-PCR SYBR Green test was successfully used in a mass-screening initiative targeting nearly 4500 asymptomatic children under the age of 12. Testing was performed at a reasonable cost of less than € 0.8 per child, and in some cases, the saliva test outperformed nasopharyngeal rapid antigen tests in identifying infected children. Whole genome sequencing revealed that the antigen testing failure could not be attributed to a specific lineage of SARS-CoV-2.

To further reduce testing costs, we produced all the necessary enzymes and established a new RT-PCR protocol based on the EvaGreen dye. Overall, this work strongly supports the view that RT-PCR saliva tests based on DNA-intercalating dyes represent a powerful strategy for community screening of SARS-CoV-2. The tests can be easily applied to other infectious agents and, therefore, constitute a powerful resource for an effective response to future pandemics.

## Introduction

One lesson from COVID-19 is that timely and accurate diagnosis, coupled with strong testing capacity and effective contact tracing, is crucial in controlling the early stages of a pandemic. In fact, COVID-19 testing played a pivotal role in providing a clear, real-time understanding of the global infection rate. This valuable information has been instrumental in guiding the implementation of measures by public health authorities [1, 2]. As a result, many countries recognized the importance of testing and made significant investments to enhance their testing capacity during that period [3, 4]. Testing primarily relied on the gold standard test for COVID-19 diagnosis, which involves real-time RT-PCR with fluorogenic probes. This test amplifies and detects the genetic material of SARS-CoV-2, which is obtained from the nasopharyngeal or oropharyngeal fluid through swab collection performed by a healthcare professional.

Although the gold standard test for COVID-19 is highly sensitive and specific, it presents several challenges. It is expensive, has long turnaround times, requires skilled personnel throughout the process, from sample collection to result delivery, and the nasopharyngeal or oropharyngeal swab can be uncomfortable. These factors limit its suitability for widespread public health testing. As a result, there has been a push to develop cheaper and faster alternative nucleic acid amplification tests (NAATs) that could reliably detect SARS-CoV-2 in clinical specimens other than nasopharyngeal or oropharyngeal swabs, such as saliva [5, 6]. Saliva offers the advantage of easy self-collection, eliminating the requirement for healthcare professionals at collection points, which not only reduces testing costs but also lowers the risk of transmission. Studies have shown that saliva performs similarly to nasopharyngeal or oropharyngeal samples in detecting SARS-CoV-2 [7–11]. Moreover, saliva can serve as a route for infection [12] and saliva from asymptomatic individuals with COVID-19 has demonstrated potential for viral transmission [12].

Several strategies have been attempted to reduce the costs associated with COVID-19 diagnosis. One promising and cost-effective approach, involves the use of isothermal amplification. This method amplifies the viral RNA at a single temperature allowing for its detection using simple procedures, without the need for complex and expensive instrumentation [13]. Another approach focuses on reducing the cost of RT-PCR-based tests. This has been achieved through the development and testing of protocols that utilize saliva, pooled samples, or DNA-intercalating dyes instead of fluorogenic probes [14–17]. DNA-intercalating dyes, such as SYBR Green, are particularly appealing due to their lower cost compared to fluorogenic probes. Additionally, assays using DNA-intercalating dyes are easier and faster to set up, and they are commonly employed in non-clinical laboratories. These features make diagnostic tests based on real-time PCR with DNA-intercalating dyes a promising strategy for decentralizing diagnosis and increasing the testing capacity and autonomy of countries during a pandemic.

Previous studies have demonstrated that singleplex or multiplex SYBR Green-based one-step or two-step RT-PCR tests designed for COVID-19 exhibit excellent sensitivity. One advantage of these tests, as opposed to RT-PCR with fluorogenic probes, is that they allow for assessment of the amplification specificity by simply examining the amplicon melting curves at the end of the PCR [18]. While most SYBR Green-based RT-PCR protocols have been established for use with RNA isolated from naso- or oropharyngeal specimens [11, 16, 19–27], there is promising evidence for the use of saliva samples with this type of tests. Ganguly *et al.* amplified synthetic SARS-CoV-2 RNA spiked into saliva specimens from healthy donors [28]. Rozanski and colleagues conducted population screening for SARS-CoV-2 infection, using a two-step protocol applied to RNA isolated from pooled saliva samples [17]. Finally, Azzi *et al.* detected SARS-CoV-2 RNA isolated from saliva samples of COVID-19 patients using a one-step RT-PCR approach [29].

In this study, we have successfully developed a one-step singleplex SYBR Green-based RT-PCR test for the accurate detection of SARS CoV-2 directly from saliva clinical samples. The test can also be applied to RNA samples isolated from naso- or oropharyngeal swabs, offering great flexibility in testing options. Our study also highlights the effectiveness and practicality of using the direct saliva test in a community setting. The results suggest that implementing this type of testing has the potential to significantly increase our testing capacity and improve our ability to manage and control infectious outbreaks more effectively in the future.

## Materials and methods

### Sample collection, processing and storage

Nasopharyngeal (NP) specimens were collected at Hospital das Forças Armadas or at the testing center of Fundição de Oeiras and processed at Laboratório de Bromatologia e Defesa Biológica (Unidade Militar Laboratorial de Defesa Biológica e Química), or at ITQB NOVA, respectively. NP samples were inactivated by incubation at 95°C for 5 min.

Saliva specimens were self-collected at the Hospital and at the testing center of Fundição de Oeiras (testing optimization), or at home (community testing) and processed at ITQB NOVA. Saliva samples (approximately 1 mL) were self-collected by spitting into sterile tubes (50 mL or 1.5 mL). In order to optimize saliva testing, nasopharyngeal (NP) swab-matched samples were collected simultaneously. Participants were instructed not to eat or drink prior to testing. After collection, saliva samples were either stored at 4°C for 2-4 days or processed immediately. The saliva specimens were inactivated at 95°C for 30 min, centrifuged at 5000g for 5 min and 200 µl of the supernatant were diluted in 10x TE (final concentration, 1x) and frozen at − 80°C until analysis. The remaining non-diluted supernatant was also frozen for storage.

### RNA extraction from clinical samples

Total viral RNA was extracted from 140 µl of deactivated samples (NP or non-diluted saliva supernatants) using Viral RNA Mini Kit (QIAGEN) and eluted in 60 µl of RNAse free water.

### SARS-CoV-2 RNA standard

SARS-CoV-2 RNA standard was prepared by amplifying the N gene from the plasmid 2019-nCoV_N_Positive Control (Integrated DNA Technologies) with a T7-promoter-containing primer (5′-TAATACGACTCACTATAGGatgtctgataatggaccccaaaa-3) and the reverse primer (5′-ttaggcctgagttgagtcagc-3′). The product was transcribed *in vitro* using HiScribe T7 High Yield RNA Synthesis Kit (NEB) following the manufacturer’s instructions. Template DNA was removed using Turbo DNase (Invitrogen) and RNA was then purified using RNeasy Mini Kit (Qiagen). Standard RNA copy numbers were calculated based on the concentration measured using Epoch Take3 spectrophotometer (Biotek).

### RT-PCR with SYBR Green

One-step qRT-PCR was performed with SYBR FAST One-Step qRT-PCR Master Mix Kit (KAPA Biosystems) according to the manufacturer’s instructions. 4 µL of sample (saliva or RNA from NP swabs) were added to the master mix to a final volume of 10 µL in a 96-well PCR plate. RT-PCR was performed in the LightCycler 480 (Roche) or the LightCycler 96 SW 1.1, using the following settings: reverse transcription step (50°C for 10 min and 95°C for 3 min), amplification step (40 or 33 cycles of 94°C for 30 s; 58°C for 30 s and 72°C for 5 s), melting curve step (95°C for 5 s; 65°C for 1 min and 97°C in continuous mode) and final cooling step (40°C, 30 s). The N gene was amplified with the primers 2019-nCoV N1-F (5’-GACCCCAAAATCAGCGAAAT-3’) and 2019-nCoV_N1-R (5’-TCTGGTTACTGCCAGTTGAATCTG-3’) [14]. The LC480 Instrument Software v1.5.0 was used to calculate the cycle threshold (CT) using the *2^nd^ derivative method* for amplification reactions with 40 cycles and the *Fit points method* for amplification reactions with 33 cycles. T_m_ calling was performed on all reactions to confirm the presence of the specific amplification product and to dismiss any false positives.

### RT-PCR with fluorogenic probes

For samples collected at Hospital das Forças Armadas, SARS-CoV-2 N gene and an internal control (RNase P) were amplified by RT-PCR using the TaqMan 2019-nCoV Assay Kit v1 (Thermofisher) with TaqMan Fast Virus 1-step Master Mix (Thermofisher) and the CFX96 thermocycler (BioRad), according to the manufacturer’s instructions For samples collected at the testing center of Fundição de Oeiras, SARS-CoV-2 ORF1ab, N gene and E gene were amplified by RT-PCR using the 2019-nCoV Real-time fluorescent RT-PCR kit (Fosun) and the LM2912 thermocycler (Fosun Diagnostics), according to the manufacturer’s instructions.

### Expression and purification of Taq DNA polymerase and MashUP reverse transcriptase (RT)

The *Thermus aquaticus* (Taq) DNA polymerase gene cloned into the pUC18 DNA vector was used to transform *E. coli* DH5α competent cells, which were then induced with 0.5 mM IPTG for 18 hours at 37 °C, 180 rpm. Cells were harvested by centrifugation at 4 °C and resuspended in a buffer solution containing 50 mM Tris-HCl pH 7.9, 50 mM glucose and 10 mM EDTA. After the addition of lysozyme (4 mg/ml), the cell suspension was incubated at room temperature for 15 min, diluted twice with a solution containing 10 mM Tris-HCl pH 7.9, 50 mM KCl, 1 mM EDTA, 0.5 % (v/v) Tween 20, 0.5 % (v/v) Nonidet P40 and further incubated at 75 °C with agitation for 1h, to precipitate *E.coli* proteins. Precipitated proteins were separated by centrifugation at 12000g for 20 minutes at 4°C and the supernatant was subjected to buffer exchange (10 mM Tris-HCl pH 8, 100 mM NaCl, 0.1 mM EDTA, 0.5 mM DTT, 1 % (v/v) Triton X-100, 50 % (v/v) glycerol). Protein aliquots were flash frozen in liquid nitrogen and stored at −80 °C until ready for use. The MashUp RT was obtained from a plasmid kindly provided by https://pipettejockey.com, according to the protocol detailed in [30].

### In-house made one-step EvaGreen-based RT-PCR assay

One step RT-PCR was performed with 1x EvaGreen dye (Biotium), the in-house made enzymes Taq DNA polymerase and MashUp RT (0.5 µl each), 1x reaction buffer (NZYTech), 2.5 mM MgCl_2_ (NZYTech), 12.5 mM KCl and 0.5 mM dNTPs (NZYTech). The N gene was amplified using 200 nM of the primer pair 2019-nCoV_N1-F and 2019-nCoV_N1-R [14]. 4 µL of RNA were added to the reaction mix to a final volume of 20 µL in a 96-well PCR plate. The analysis was performed in the LightCycler 480 (Roche) using the following settings: reverse transcription step (50°C for 10 min and 95°C for 3 min), amplification step (33 cycles of 94°C for 30 s, 60°C for 30 s and 72°C for 5 s), melting curve (95°C for 5 s; 65°C for 1 min and 97°C in continuous mode) and final cooling step (40°C, 30 s). CT determination and Tm calling was performed following the methods described above.

### Vectors and primer design for VOC detection

Two plasmids were synthesized by GenScript, one containing the sequence of the original SARS-CoV-2 strain (pWT), and other carrying the specific mutation Δ3675-3677 in ORF1a (pVOCα). One-step RT-PCR was performed with SYBR FAST One-Step qRT-PCR Master Mix Kit (KAPA Biosystems) according to the manufacturer’s instructions. 4 µL of each DNA plasmid (pWT or pVOCα) were added to the master mix to a final volume of 10 µL in a 96-well PCR plate. The forward (Fw_Orf_mis 5’-GGTTGGATATGGTTGATACTAGTTTGCA-3’) and reverse (Rv_Orf 5’-GTTCTTGCTGTCATAAGGATTAGTAAC-3’) primers were used at a final concentration of 100 nM. The analysis was performed in the LightCycler 480 (Roche) using the following settings: reverse transcription step (50°C, 10 min and 95°C, 3 min), amplification step (35 cycles of 94°C, 30 s; 62°C, 30 s and 72°C, 5 s), melting curve step (95°C for 5 s; 65°C for 1 min and 97°C in continuous mode) and a final cooling step (40°C, 30 s). CT determination and Tm calling was performed following the methods described above. The two RNA samples analysed were obtained from patients who had confirmed infections with the SARS-CoV-2 variants Alpha and Omicron BA.1. Variant identification was previously confirmed through whole genome sequencing, using long reads from Oxford Nanopore Technologies (see below).

### Community screening

From January 2022 to March 2022, in partnership with the Municipality of Oeiras, Portugal, the ITQB NOVA provided free saliva testing using the SYBR Green-based one-step RT-PCR to volunteer students (aged 3 to 11 years) from all public schools in Oeiras. Circa 9500 saliva collection kits were provided and 4445 tests were completed. Students were instructed to collect the saliva sample in the morning on a pre-defined day before eating, drinking and brushing teeth. The samples were kept at 4°C, promptly transported to ITQB NOVA and analysed on the same day of collection. Confirmatory standard RT-PCR tests with NP sampling were performed on participants with positive saliva tests, typically 24-48 hours after the notification of the result. Students who had tested positive for COVID-19 in the 3 months prior to the screening initiative were excluded from the study.

### Genome sequencing and bioinformatics analysis

RNA samples were directly used for genome sequencing following the classic PCR tilling of SARS-CoV-2 virus protocol, from Oxford Nanopore Technologies, and further described in the ARTIC network protocol (https://artic.network/ncov-2019; https://www.protocols.io/view/ncov-2019-sequencing-protocol-bbmuik6w). This constitutes an amplicon-based whole-genome amplification strategy using tiled, multiplexed primers [31]. In brief, SARS-CoV-2 gRNA was reverse transcribed and the obtained cDNA was amplified (400bp amplicons), recurring to two separate pools of tiling primers (ARTIC V3), using the NEBNext ARTIC SARS-CoV-2 Companion Kit (Oxford Nanopore Technologies, Oxford, UK). The generated pools of multiplexed amplicons were combined, for each sample, purified and prepared for barcoding (Native Barcoding Expansion, EXP-NBD104), to allow multiplex sequencing. Additionally, sequencing adapters were ligated to the pooled barcoded samples, according to manufacturers’ instructions. Sequencing libraries were sequenced in multiplex on one flow cell (R9.4.1) using a MinION Mk1C device, according to the settings described in the classic PCR tilling of SARS-CoV-2 virus protocol. MinKNOW (v21.11.) software was used for basecalling (min Qscore 8) and reads demultiplexing, so downstream bioinformatics analysis could be performed. All bioinformatics analysis were performed using EPI2ME (v3.4.2) and the Fastq QC + ARTIC + NextClade workflow. A consensus sequence was obtained using the genome sequence of SARS-CoV-2 Wuhan-Hu-1/2019 virus (GenBank accession number MN908947) as reference [32]. All the SARS-CoV-2 genome sequences included in the analysis had >70% of the genome covered by at least 20-fold, presenting an overall quality score of 11.5; regions with a depth of coverage below this threshold were automatically marked as ambiguous and undefined bases “N” were introduced in the consensus sequence. Clade and lineage assignments were performed using Nextclade (v2.4.1) (https://clades.nextstrain.org/) (accessed on 04 August 2022).

#### Ethics statement

Ethical approval was given by the Institutional Review Board of Hospital das Forças Armadas (HFA), Portugal, (samples collected at HFA) or by the Instituto de Higiene e Medicina Tropical – Instituto de Tecnologia Química e Biológica (ITQB NOVA) Ethics Committee (samples collected at Fundição de Oeiras), Portugal. The study was conducted in accordance with the European Statements for Good Clinical Practice and the Declaration of Helsinki of the World Health Medical Association. Written informed consent was obtained from all participants.

## Results

### Sensitivity and specificity of a SYBR Green-based one-step RT-PCR assay in detecting SARS-CoV-2 in nasopharyngeal samples

The costs associated with the reference diagnostic test for COVID-19 have made it unaffordable for countries with limited resources and inadequate for periodic mass screening purposes. The interest in overcoming such limitation prompted us to test an alternative RT-PCR protocol that uses inexpensive DNA-intercalating dyes instead of costly fluorogenic probes. After inspecting the RT-PCR kits using intercalating dyes available in the market, we opted for the SYBR FAST One-Step RT-PCR Master Mix Kit (KAPA Biosystems) due to its competitive price. For the PCR reaction we chose a singleplex strategy to amplify a fragment of the viral RNA (N gene) using the N1 primer set designed and proved to be specific for SARS-CoV-2 by other authors [14]. We first evaluated the analytical sensitivity of the assay by testing tenfold serial dilutions of the *in vitro* transcribed SARS-CoV-2 N gene (IVT N-gene) in order to estimate the minimum number of genomic copies of the virus that could be detected. The limit of detection (LoD) of the assay was found to be between 1 and 10 copies per reaction (Fig. 1A). However, analysis of the amplification and amplicon melting temperature curves indicated the presence of unspecific amplifications in the no template control (NTC) after 40 PCR cycles (Fig. 1B). This was likely due to the formation of primer dimers, which could interfere with the accurate identification of positive samples. In order to mitigate this issue, we adjusted the number of PCR cycles to 33. We reasoned that while this adjustment may slightly decrease the sensitivity of the assay, it would greatly enhance the specificity. With this modified setup, we successfully eliminated non-specific amplifications, and we were able to reliably detect as few as 25 copies of the IVT N-gene. (Fig. S1 and Fig. 1C).

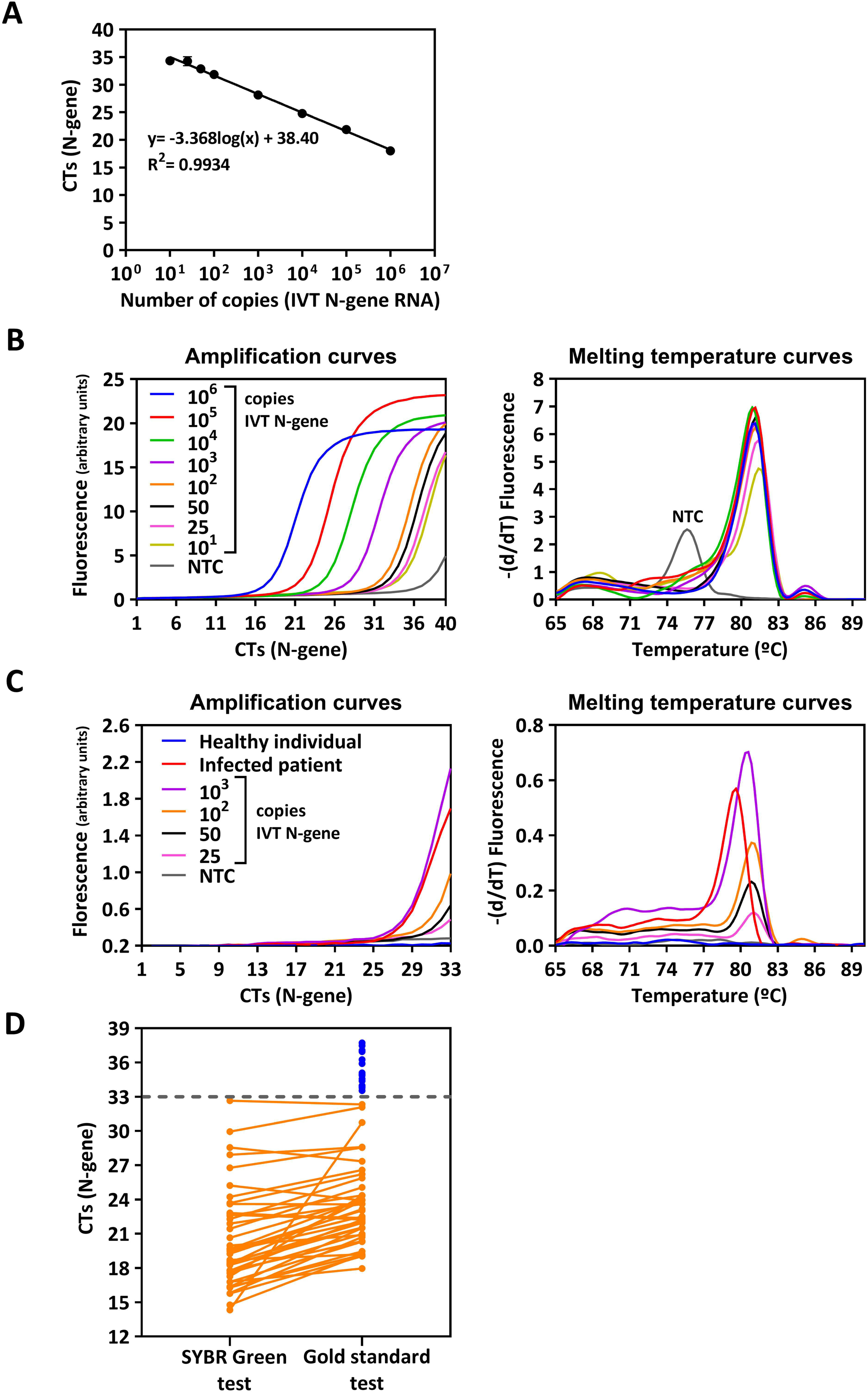

We then used a set of 104 NP clinical samples to evaluate whether the SYBR Green-based RT-PCR could accurately detect SARS-CoV-2 in the NP fluid of infected individuals. The test was benchmarked against the gold standard for COVID-19 diagnosis (RT-PCR with fluorogenic hybridization probes), using RNA isolated from those NP samples. For the majority of positive samples, we observed a close correlation between the cycle threshold (CT) values obtained from both the SYBR Green-based test and the gold standard test (Fig. 1D). For samples with CTs ≤ 33, the SYBR Green test showed excellent agreement (100%) with the gold standard. However, for samples with CT values greater than or equal to 33 (15 samples), the SYBR Green assay failed to identify them as positive cases (Fig. 1D). Samples that were determined to be negative by the gold standard test also yielded negative results in the SYBR Green assay when subjected to 33 cycles of amplification. This indicates a high specificity of the SYBR Green assay under these conditions. It is important to note that when the amplification was extended to 40 PCR cycles, non-specific amplification products were observed, leading to the identification of false positive results. This further supports our decision to set the number of PCR cycles to 33 to minimize the occurrence of false positives.

Overall, these results indicate that the SYBR Green-based one-step RT-PCR assay reliably detects medium to high viral loads in RNA isolated from NP swabs.

### Analytical sensitivity of an in-house made one-step EvaGreen-based RT-PCR

During the initial months of the COVID-19 pandemic, all countries experienced testing slowdowns due to a scarcity of reagents required for the gold standard test, which was the only available COVID-19 diagnostic test at that time. This global experience has emphasized the importance of each nation’s pandemic preparedness including self-sufficient production of testing reagents, independent of specific commercial suppliers. In a previous study, we presented a protocol for producing a reverse transcriptase, which we used in an in-house made RT-LAMP assay tailored for COVID-19 [30]. Here we have combined this enzyme with a newly produced Taq DNA polymerase and established an in-house made one-step RT-PCR test for detecting SARS-CoV-2. Following an optimization process involving fine-tuning of several reaction parameters (Fig. S2), we decided to utilize the intercalating EvaGreen dye for the purpose of detection.

The LoD of the RT-PCR EvaGreen assay was determined as described above and found to fall in the range between 100 and 1000 copies of viral RNA (Fig. 2A). Under the defined conditions, no unspecific amplifications were observed (Fig. 2B). Although the test exhibited lower sensitivity than the SYBR Green-based RT-PCR test, it reliably detected the genetic material of SARS-CoV-2 in RNA samples isolated from NP swabs, as depicted in Fig. 2C.

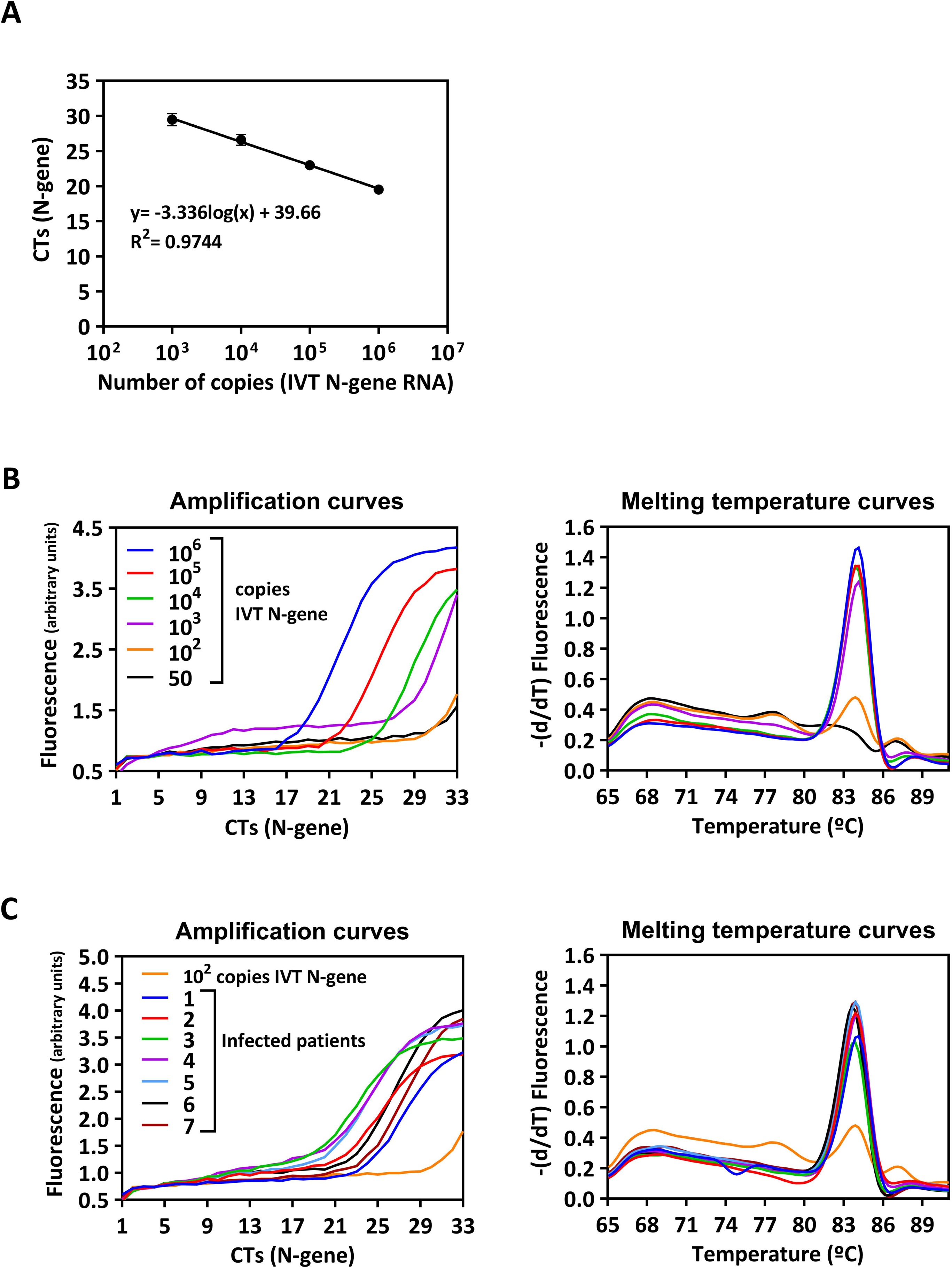

### The SYBR Green one-step RT-PCR can be used to detect variants of concern

In late 2020, the emergence of SARS-CoV-2 variants that posed an increased threat to public health [33, 34] urged global monitoring of variants of concern (VOCs) in order to control their spread, provide information to decision-makers, and implement appropriate measures. Mass testing plays a crucial role in this scenario. However, VOC identification primarily relies on expensive genome sequencing methods, which not only add to the economic burden imposed by the pandemic but also restrict VOC identification in low-resource settings. Therefore, we conducted experiments to determine whether our simple and cost-effective SYBR Green-based RT-PCR assay could be utilized for VOC detection. We employed a strategy that targets fingerprint mutations specific to those variants. To demonstrate the proof-of-concept, our focus was on a specific region of the ORF1a gene of SARS-CoV-2 that exhibited a deletion in the Alpha variant’s genome (Δ3675-3677) [35]. Inspired by the allele specific PCR method [36], we designed an oligonucleotide primer (Fw_Orf_mis, Fig 3A) whose 3’ end perfectly matched the sequence of the Alpha variant, but not that of the original wild-type strain (Wuhan-Hu1 strain, WT) due to the Δ3675-3677 deletion in the former. To further strengthen variant-specific primer hybridization, an additional mismatch was introduced at the 3’ end of the primer (Fig. 3A).

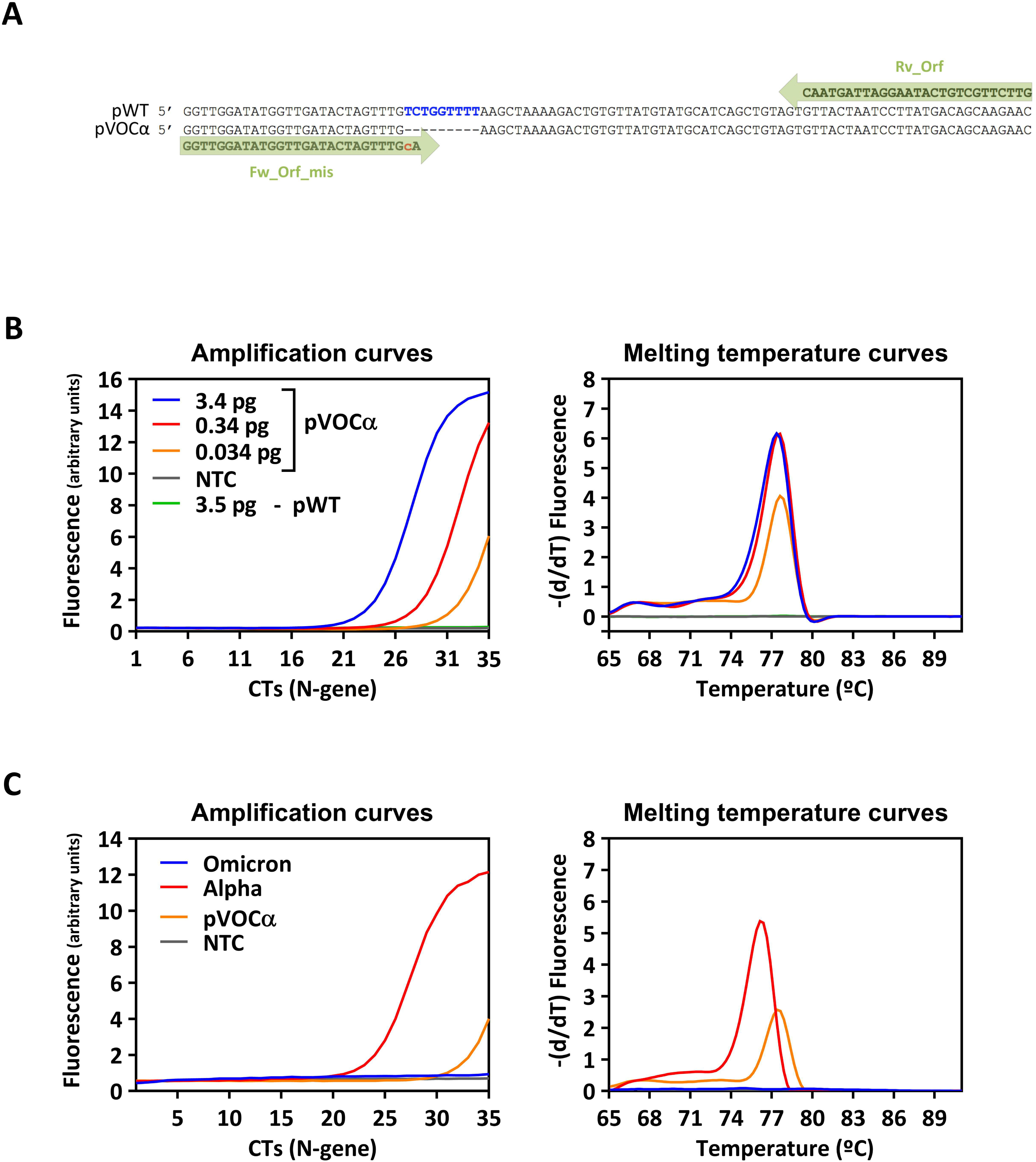

We next constructed two plasmids, pVOCα and pWT, each carrying a fragment of the ORF1a region with or without the deletion Δ3675-3677, respectively. Each plasmid was used as a DNA template in the SYBR Green-based RT-PCR assay to evaluate its capacity to distinguish different SARS-CoV-2 variants. Amplification conditions were first optimized by testing several primer concentrations, different annealing temperatures, and varying the number of amplification cycles (data not shown). After determining the optimal conditions, we performed amplification on serial dilutions of pVOCα and pWT. Notably, robust detection was achieved for all dilutions of pVOCα, while pWT remained undetectable (Fig. 3B). Lastly, we assessed the performance of the assay using two real-life RNA samples isolated from patients infected with the SARS-CoV-2 variants Alpha and Omicron BA.1. As expected, amplification was observed only in the sample containing the Alpha variant (Fig. 3C) since the specific deletion (Δ3675-3677) targeted by the assay is absent in Omicron BA.1 [37].

### The SYBR Green-based one-step RT-PCR test demonstrates great analytical and clinical sensitivity in directly detecting SARS-CoV-2 from saliva samples

The advantages of using saliva samples instead of NP swabs for diagnosing COVID-19 have prompted us to investigate whether the one-step SYBR Green-based assay could be employed for identifying SARS-CoV-2 in saliva without the need for prior RNA. Apart from offering a simpler and less invasive sample collection procedure, such a test would expedite the time to results and further reduce diagnostic costs. Prior to amplification, the samples underwent a straightforward pre-treatment process, which involved viral inactivation by heating at 95°C for 30 min and addition of TE buffer. Importantly, the tubes containing the saliva samples remained sealed following self-collection and throughout the entire inactivation process, ensuring the safety of the operating personnel.

The analytical sensitivity of the SYBR Green-based RT-PCR saliva test was assessed by spiking saliva samples obtained from healthy donors (confirmed by the gold standard) with different amounts of the IVT N-gene. As described above, to decrease nonspecific amplification we defined a threshold of 33 cycles in the RT-PCR reaction. The test was able to detect as low as 10 copies of viral RNA per reaction (Fig. 4A), and no unspecific products were observed (Fig. 4B). The analysis of the amplification and melting temperature curves (Fig. 4B) revealed clear profiles, and the protocol proved to be efficient in directly detecting SARS-CoV-2 from saliva samples of infected individuals (Fig. 4C). We also evaluated the sensitivity and specificity of the saliva test in detecting the genetic material of SARS-CoV-2 from clinical samples. We conducted an analysis on paired NP swab and saliva samples from 129 individuals using both the gold standard RT-PCR (NP swab) and the SYBR Green-based RT-PCR (saliva) tests, with or without prior RNA extraction, respectively. According to the gold standard, 29 samples were positive and 100 were negative for SARS-CoV-2 infection, with the CTs of the former varying between 18.3 and 26.6. The SYBR Green-based RT-PCR successfully detected viral genetic material directly from the saliva samples, i.e., without the need for RNA pre-extraction, in all but one positive sample (Fig. 4D). All 100 samples scored as negative by the gold standard test were also identified as negative by the SYBR Green-based RT-PCR, demonstrating the excellent specificity of the saliva test (Table 1). By eliminating the RNA extraction step, the total assay time was reduced to 100 minutes, and the estimated cost per sample was less than € 0.8, offering significant time and cost savings (Table 1).

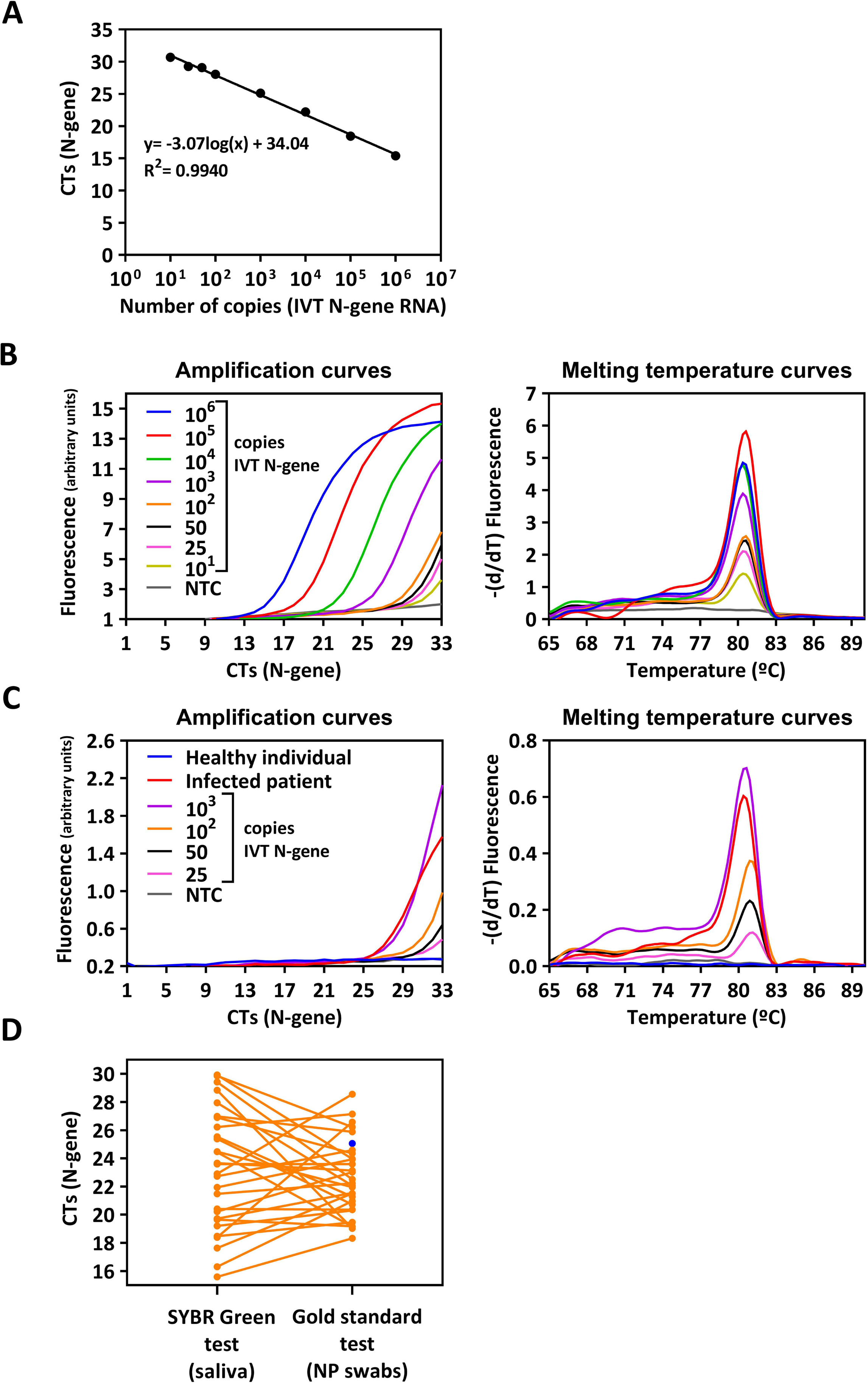

### COVID-19 mass-testing using the SYBR-Green RT-PCR saliva test

To assess the reliability and usefulness of the SYBR-Green RT-PCR saliva test for community mass screening, we made the test available to students attending public schools in Oeiras (Lisbon district). The target population consisted of children aged 3-12 who had recently become eligible for COVID-19 vaccination (which was ongoing at the time of sampling) and were not being included in the regular testing protocols followed by Portuguese public schools during that period. Over the course of one month, a total of 4445 students underwent testing using the SYBR-Green RT-PCR saliva test, leading to the identification of 80 asymptomatic children whose saliva samples tested positive for SARS-CoV-2 infection. The gold standard test confirmed the infection in all students who had received a positive result from the saliva test. No false positives were reported.

Interestingly, among the 80 children who tested positive with our saliva test, five tested negative when using pharmacy rapid antigen tests (RATs) with NP sampling. To determine if specific lineages or variants of SARS-CoV-2 could be responsible for the observed differences in test outcomes, we conducted viral genomic sequencing on the RNA extracted from the positive saliva samples. The sequencing analysis was performed on samples where the SYBR-Green RT-PCR results agreed with the RATs (three samples), as well as on samples where they disagreed (three samples). Genomic sequences were then compared with the genomic diversity of SARS-CoV-2 in Portugal (NextClade database, as of July 2022) to infer phylogenetic relationships. We found that the three viral genomic sequences retrieved from saliva samples of individuals with disparate results between the saliva and antigen tests belonged to sub-variant BA.1 (n=3/3 samples). However, this same sub-variant was identified in samples from infected individuals whose saliva test and antigen tests agreed (n=1/3 samples) (Fig. 5). This analysis ruled out the hypothesis that a specific lineage or variant was escaping antigen tests.

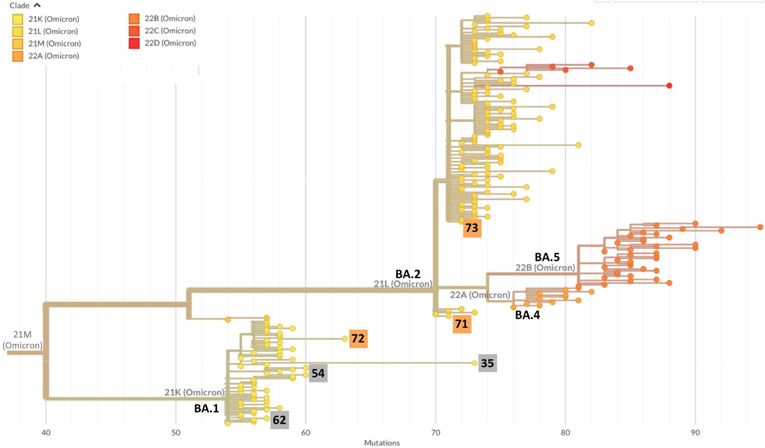

## Discussion

The COVID-19 pandemic has raised awareness of the significant importance of diagnosis in assisting the implementation of effective public health measures. However, the gold standard test for COVID-19 is complex and costly, creating challenges for many low- and middle-income countries. This disparity, coupled with limitations in the supply chain of reagents required for the gold standard test, has spurred the development of simpler and more affordable testing options.

Here we have developed an alternative RT-PCR test based on SYBR Green that demonstrates reliable performance in detecting medium to high viral loads. The test can be conducted directly on saliva specimens or RNA purified from NP swabs. While it is less sensitive than the gold standard test, it exhibits exceptional specificity, making it valuable in scenarios beyond diagnosis. In a pandemic setting, where different testing purposes exist (diagnosis, surveillance, screening), tests with lower sensitivity or specificity can still play a crucial role [38, 39]. This notion is supported by the successful implementation of public health screenings using less sensitive tests, such as rapid antigen tests (RATs) [4]. Indeed, the primary goal of public health screenings is not merely to diagnose individuals but to identify those at higher risk of transmitting the infection, typically those with high viral loads, in order to prevent outbreaks and uncontrolled spread of the disease. This is especially important for COVID-19, as the virus can be transmitted by presymptomatic or asymptomatic individuals, making symptom-based distancing less effective [40].

In screening scenarios, the speed and frequency of testing are more important than test sensitivity [2, 41]. The cost per test is, consequently, a crucial factor, and our highly specific SYBR Green-based RT-PCR saliva test, costing less than € 0.8, offers an interesting alternative to the gold standard. Furthermore, its different chemistry and reagents alleviate the strain on the supply chains associated with the gold standard test, ensuring the continuity of diagnosis.

To further reduce testing costs, we have developed a homemade one-step RT-PCR protocol based on EvaGreen. Although its sensitivity is currently inadequate for diagnosis or mass screening purposes, it can serve as a backup option for standard molecular diagnostics in case of supply chain shortages, and it provides a starting point for further optimization.

In our community screening study, the SYBR Green-based direct saliva RT-PCR test demonstrated robust performance. By eliminating the laborious RNA extraction step, the test reduced costs and accelerated result delivery. The test exhibited excellent clinical specificity, correctly identifying 80 infected children confirmed by the gold standard test. Although targeting several genes has been shown to decrease the ratio of false positives [26], using our simple singleplex assay no false positives were observed despite the considerable size of the tested population (4445 tests performed). Interestingly, our test outperformed RATs, as the latter failed to detect 5 positive samples. Whole genome sequencing analysis revealed that different SARS-CoV-2 lineages did not account for these discrepancies, suggesting that inefficient nasopharyngeal sampling or lower sensitivity of the RATs used by some pharmacies at the time may explain the differences. During the COVID-19 pandemic, multiple variants of the original SARS-CoV-2 strain have emerged as a result of acquired mutations in various regions of the viral genome. Some of these variants were initially identified due to an inability to amplify specific genomic regions using commercial primers, and subsequent confirmation was achieved through whole-genome sequencing [42]. Although not all mutations impact the progression of the pandemic, certain mutations have contributed to the emergence of more transmissible or immune-evading variants [43]. Given the significance of these variants, detecting and monitoring them in the community can be valuable for guiding public health interventions. To the best of our knowledge, there is only one published report where SARS-CoV-2 variant detection was attempted using SYBR-Green-based RT-PCR [44]. The authors used a two-step SYBR-Green RT-PCR protocol with primers targeting the spike and nsp6 gene regions, specifically focusing on the amino acid residue deletions 69/70 and 106/107/108, respectively. In our study, we attempted a similar one-step approach to detect the Δ3675-3677 mutation in the ORF1a region (data not shown), however, in our experiments, we found that to completely eliminate amplification of the original SARS-CoV-2 strain, we needed to employ the allele specific amplification strategy. Unlike the aforementioned authors, we optimized the SYBR-Green-based RT-PCR protocol for variant discrimination using plasmids (pWT and pVOC), which could simulate higher viral loads and facilitate the detection of less efficient RT-PCR reactions.

Respiratory viruses have historically been a frequent cause of pandemics [45], and the detection of these virus in saliva has emerged as valuable diagnostic tool [46–51]. In this regard, our saliva test, which can be easily adjusted for other pathogens and is capable of reliably identify (pre-)asymptomatic individuals in community scenarios, certainly offers many benefits in the battle against future silent pandemics.

## Supporting information

Supplemental images

## Data Availability

All data produced in the present work are contained in the manuscript

## Acknowledgements

We express our gratitude to the members of the COVID-19 task force of ITQB NOVA for their valuable discussions and suggestions. Specifically, we extend our thanks to Prof. Claudio M. Soares for his unwavering support. We are grateful to Dr. Alexandra Simões and Dr. Sónia Almeida for their work in conducting the RT-PCR (gold standard) analyses. We deeply thank Professor Luis Netto and Chuck Farah from the São Paulo University, Brazil, for the *Thermus aquaticus* DNA polymerase gene and the pGTf2 plasmid, respectively. We are also deeply grateful to all the members of the ITQB NOVA Saliva Testing Initiative for their invaluable support in community screening. We would like to acknowledge the extraordinary efforts of Dr. Renata Ramalho and her team, who went above and beyond their communication duties. Our heartfelt thanks go to the Oeiras Municipality, particularly Dr. Pedro Patacho, Dr. Maria Paula Rodrigues, Dr. Elisabete Brigadeiro, Marta Filipa Sousa, Dr. Luís Afonso, and Dr. Ana Alexandra Reis, without whom the mass-screening project would not have been possible. We extend our appreciation to all the Directors and Coordinators of the schools in Oeiras for their enthusiastic participation in the mass screening endeavor. Lastly, we would like to express our gratitude to the Ethical Review Board IHMT–ITQB, specifically Prof. Cláudia Conceição, for promptly addressing all our queries and providing insightful reflections on topics we had never ventured before.

This work was supported by (i) Project LISBOA-01-0145-FEDER-007660 (“Microbiologia Molecular, Estrutural e Celular”) funded by FEDER funds through COMPETE2020— “Programa Operacional Competitividade e Internacionalização” (POCI), (ii) “Fundação para a Ciência e a Tecnologia” (FCT) through the project DETECT Ref 433_613549914 (20/7/153), under the scope of the 2nd edition of the programme RESEARCH4COVID19, (iii) the project “STOP-COVID—Strategies to prevent COVID-19 by early detection of asymptomatic carriers at increased risk: epidemiological studies and validation of a rapid in-house diagnostic test”, Ref 072559, funded by FEDER “Fundo Europeu de Desenvolvimento Regional” from “Programa Operacional Regional Lisboa” and (iv) the Municipality of Oeiras

