## Supplemental images for "A low-cost molecular test for SARS-CoV-2 detection suitable for variant discrimination and community testing using saliva"

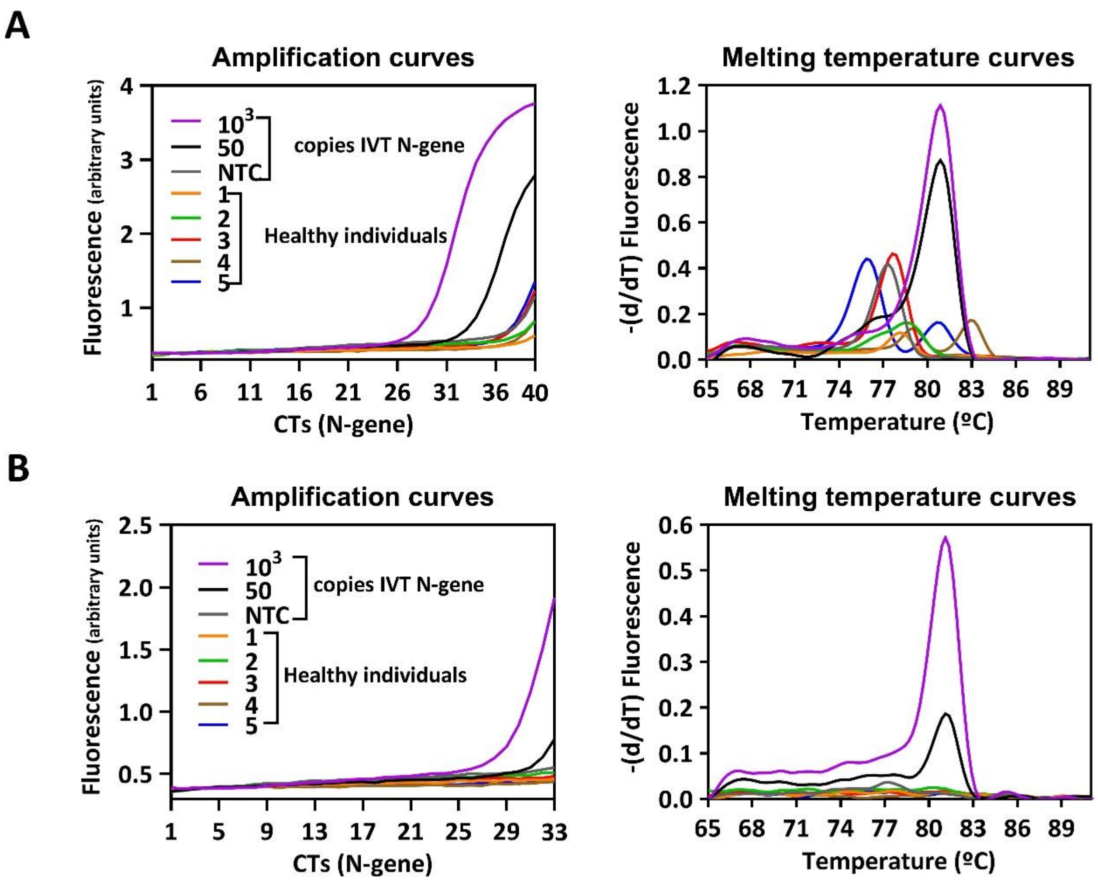


**Figure S1.** Amplification and melting curve profiles after 40 and 33 amplification cycles using the SYBR Green RT-PCR assay. Plots were obtained after **(A)** 40 cycles and **(B)** 33 cycles of amplification of SARS CoV-2 IVT N-gene and RNA extracted from NP swabs obtained from healthy individuals.

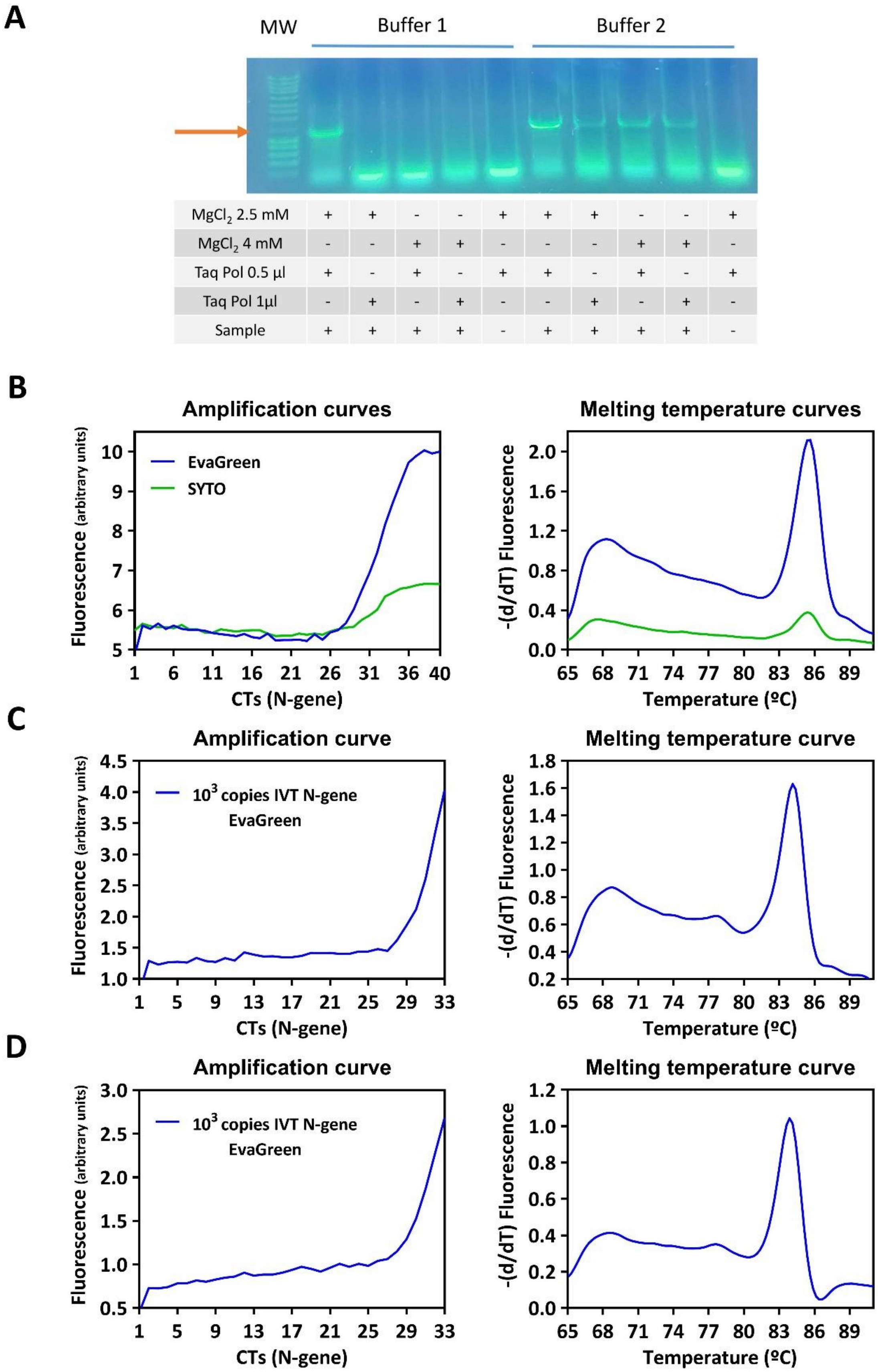
A
MW
Buffer 1
Buffer 2
MgCl2 2.5 mM + + - - + + + - - +
MgCl2 4 mM - - + + - - - + + -
Taq Pol 0.5 µl + - + - + + - + - +
Taq Pol 1µl - + - + - - + - + -
Sample + + + + - + + + + -
B
Amplification curves
Fluorescence (arbitrary units)
EvaGreen
SYTO
CTs (N-gene)
Melting temperature curves
-(d/dT) Fluorescence
Temperature (ºC)
C
Amplification curve
10^3 copies IVT N-gene
EvaGreen
CTs (N-gene)
Melting temperature curve
Temperature (ºC)
D
Amplification curve
10^3 copies IVT N-gene
EvaGreen
CTs (N-gene)
Melting temperature curve
Temperature (ºC)

**Figure S2.** Optimization of an in-house made RT-PCR assay using DNA-intercalating dyes. (**A**) Optimization of the PCR reaction conditions using the in-house produced Taq DNA polymerase. Buffer 1 and buffer 2 are two commercial buffers, from Promega (GoTaq polymerase) and NZYTech (NZYTaq II polymerase), respectively. MW, molecular weight marker (NZY DNA Ladder III). (**B**) Amplification and melting curves of $10^3$ copies of IVT N-gene using two intercalating dyes, EvaGreen (Biotium) and SYTO (ThermoFisher). The analysis was performed in the LightCycler 480 (Roche) using the following settings: reverse transcription step (50ºC, 10 min and 95ºC, 3 min), amplification step (40 cycles of 94ºC, 30 s; 58ºC, 30 s and 72ºC, 5 s), melting curve step (95ºC for 5 s; 65ºC for 1 min and 97ºC in continuous mode) and a final cooling step (40ºC, 30 s). (**C**) Amplification and melting curve of $10^3$ copies of IVT N-gene with EvaGreen. The analysis was performed in the LightCycler 480 (Roche) using the following settings: reverse transcription step (50ºC, 10 min and 95ºC, 3 min), amplification step (33 cycles of 94ºC, 30 s; 58ºC, 30 s and 72ºC, 5 s), melting curve step (95ºC for 5 s; 65ºC for 1 min and 97ºC in continuous mode) and a final cooling step (40ºC, 30 s). (**D**) Amplification and melting curve of $10^3$ copies of IVT N-gene with EvaGreen. The analysis was performed in the LightCycler 480 (Roche) using the following settings: reverse transcription step (50ºC, 10 min and 95ºC, 3 min), amplification step (33 cycles of 94ºC, 30 s; 60ºC, 30 s and 72ºC, 5 s), melting curve step (95ºC for 5 s; 65ºC for 1 min and 97ºC in continuous mode) and a final cooling step (40ºC, 30 s).
